# Long-term exposure to air pollution and risk of adult-onset asthma and COPD: Danish nationwide cohort study

**DOI:** 10.1101/2025.06.11.25328651

**Authors:** Rina So, Jiawei Zhang, Marie Bergmann, Youn-Hee Lim, Gonzalo Hevia-Ramos, George Napolitano, Ebba Malmqvist, Anna Oudin, Kajsa Pira, Cale Lawlor, Tanya Andersson Nystedt, Charlotte Suppli Ulrik, Jie Chen, Stéphane Tuffier, Thomas Cole-Hunter, Steffen Loft, Lau Caspar Thygesen, Massimo Stafoggia, Kees de Hoogh, Evangelia Samoli, Bert Brunekreef, Gerard Hoek, Zorana J. Andersen

## Abstract

**Background:** We investigated the association of long-term exposure to fine particulate matter (PM_2.5_), black carbon (BC), and nitrogen dioxide (NO_2_) with incidence of adult-onset asthma and chronic obstructive pulmonary disease (COPD) in a Danish nationwide cohort.

**Materials and methods:** We followed 3.2 million Danish residents aged 30 years or older on January 1^st^, 2000 until December 31^st^, 2018, for: (1) incidence of asthma (first hospital contact); (2) incidence of COPD (first hospital contact), and; (3) combined incidence of asthma or COPD (first prescription for obstructive airway disease (OAD) medication). Annual mean concentrations of air pollutants were estimated using European-wide hybrid land-use regression models. Cox proportional hazard models were used to investigate the association of air pollutants with the three outcomes, adjusting directly for age, sex, and socioeconomic status, and indirectly for smoking and body mass index.

**Results:** Over a mean follow-up of 16 years, 52,648 participants developed asthma, 146,269 developed COPD, and 393,211 were prescribed OAD medication. We found positive, statistically significant associations of PM_2.5_, BC, and NO_2_ with all outcomes, strongest for asthma and weakest for OAD medication. The most robust associations were seen for BC in two-pollutant models.

**Conclusion:** In a large Danish nationwide analysis, air pollution (PM_2.5_, BC, and NO_2_) is an important predictor for adult-onset asthma and COPD, with the strongest associations for asthma. Our findings present novel evidence highlighting the relevance of BC as an independent risk factor for asthma and COPD, beyond the effects of PM_2.5_.

**What is already known on this topic:** Long-term exposure to fine particulate matter (PM_2.5_) and nitrogen dioxide (NO_2_) is linked to the increased risk of adultlJonset asthma and COPD, as supported by WHO reviews and meta-analyses of European cohorts. Black carbon (BC), a trafficlJ and combustionlJrelated component of PM_2.5_, is recognized for its toxicological effects on inducing inflammation and oxidative stress in the airway. Still, epidemiological evidence on its role in incident asthma and COPD is scarce and inconclusive, and very few directly contrasted BC and PM_₂_.lJ effects in twolJpollutant models.

**What this study adds:** In a nationwide Danish cohort of >3 million adults with a mean 16 years’ follow-up, long-term exposure to BC showed the strongest, most robust associations with first hospital contact for asthma and COPD, and with first prescriptions for obstructive airway disease medications, even after adjusting for PM_2.5._

**How this study might affect research, practice or policy:** These results underscore BC as an independent risk factor, supporting its inclusion in air-quality regulations and guiding efforts to target combustion-related pollutants for respiratory disease prevention.

## INTRODUCTION

Air pollution accounted for 8.1 million deaths globally in 2021, becoming the second leading risk factor for death after high blood pressure [1]. The World Health Organization (WHO) Air Quality Guideline stated that despite some improvements in air quality globally, almost everyone is still breathing air that is harmful to health, especially particulate matter with a diameter < 2.5 µm (PM_2.5_) exceeding 5 µg/m^3^ [2]. The burden is especially high for chronic lung diseases, as inhaled PM_2.5_ can cause inflammation, sensitization, and oxidative stress in the lung tissue, causing airway damage and impaired lung function [3], the key pathologies in asthma and COPD [4]. It is estimated that 48% of COPD mortality globally is attributed to air pollution [2]. Furthermore, the recent WHO project ‘Estimation of Morbidity from Air Pollution and its Economic Costs’ [5] concluded that there was sufficient evidence to support a causal relationship of long-term exposure to nitrogen dioxide (NO_2_) with adult-onset asthma[6], and PM_2.5_ with COPD incidence[7]. A recent meta-analysis of nine studies on long-term exposure to air pollution and adult-onset asthma showed significant associations with PM_2.5_ and NO_2_ [8].

However, there are important knowledge gaps related to air pollution and chronic respiratory diseases that remain poorly understood. The WHO Air Quality Guideline has identified black carbon (BC) as a pollutant of ‘emerging concern’ for which there was not enough evidence to set guidelines in 2021, but toxicological evidence showed its adverse impacts on health (inflammation, oxidative stress, carcinogenic effect). BC is not a regulated pollutant (unlike PM_2.5_ and NO_2_) and therefore is not routinely monitored, resulting in a lack of reliable exposure data for epidemiological studies, which is necessary to provide evidence on health effects to, in turn, guide regulation. BC, a component of PM_2.5_, is formed from the incomplete combustion of biomass and fossil fuels from various sources, including on- and off-road diesel engines, coal and wood burning for residential heating and cooking, coal power plants, and the open burning of agricultural fields, among others. BC is also a strong climate change forcer, as its black surface absorbs energy, converting it to heat and contributing to global warming. More evidence on the health effects of BC is urgently needed to support the EU

Green Deal Zero Pollution Ambition, which aims to reduce the health impacts of air pollution by half by 2050, as well as efforts to mitigate climate change. Epidemiological evidence on BC and chronic lung disease is limited. A recent meta-analysis revealed associations between long-term exposure to BC and respiratory mortality, based on nine studies; however, it identified only three studies on COPD mortality, yielding inconclusive findings [9]. There are only two studies on long-term exposure to BC and asthma incidence [10, 11] and four on COPD incidence [12–15]. The large European Study of Cohorts for Air Pollution Effects (ESCAPE) found no association of BC with asthma [11] and weak associations with COPD [13], whereas the more recent study from the Effects of Low-Level Air Pollution: A Study in Europe (ELAPSE) detected associations of BC with both outcomes [10, 12].

Furthermore, both a smaller Nordic RHINE study [15] and a large Vancouver administrative cohort [14] reported associations of BC with COPD incidence. To help guide BC regulation, it is crucial to understand whether the association of BC with asthma or COPD is independent from that of PM_2.5_ in two-pollutant models. Only two studies have done this [10, 12], suggesting BC associations with asthma and COPD are independent of PM_2.5_, and more studies are warranted to confirm or refute this.

In the present study, we examine the association between long-term exposure to PM_2.5_, BC, and NO_2_ and the incidence of adult-onset asthma and COPD in a nationwide Danish cohort, applying two definitions of asthma and COPD: one based on hospital contact data and the other on filled medication prescriptions from unique Danish national registers.

## METHODS

### Study population

We used a Danish nationwide administrative cohort, comprising all Danish residents aged 30 years or older (N = 3,323,612) as of January 1, 2000 (baseline), who had resided in Denmark for at least one year prior, as defined in a previous study [16]. We obtained sex, age, individual-level socioeconomic status (SES), including household income, educational level, and occupational and marital status, at baseline, and parish- and regional-level SES (mean household income and unemployment rate) for 2001. The region was assigned based on individuals’ baseline residential addresses.

### Outcome definition

To define the incidence of asthma and COPD based on hospital contacts, we linked individuals to the Danish National Patient Registry [17] by the personal identification numbers. This registry includes information on all hospital contacts, including inpatient (since 1977), outpatient (since 1995), and emergency room visits (since 1995) in Denmark. The incidence was defined as the first hospital contact between baseline and the end of follow-up with primary asthma or COPD discharge diagnosis, excluding individuals with hospital contact for asthma or COPD, respectively, before baseline (prevalent cases). The following International Classification of Disease 8^th^ version (ICD-8; before 1994) and 10^th^ version (ICD-10; from 1994) were used to extract the diagnosis of asthma and COPD: ICD-8: 493 or ICD-10: J45-J46 for asthma, and ICD-8: 490-492 or ICD-10: J40-44 for COPD.

In addition, we defined the incidence of asthma and COPD based on redeemed prescriptions for medications used to treat obstructive airway diseases (OAD). As OAD medication is prescribed for both asthma and COPD, it is not possible to reliably distinguish between the incidence of asthma and COPD based on OAD medication prescriptions; thus, the incidence of OAD is a combined incidence of asthma and COPD. The individuals were linked to the Danish National Prescription Registry [18], which includes information on all redeemed prescriptions in Denmark since 1995. Individuals were considered as incident cases of asthma or COPD if they had at least two prescriptions for OAD (ATC-code: R03) within 12 months between baseline and the end of follow-up, with the incidence date being the date of the second filled OAD prescription, after excluding individuals with OAD prescriptions before baseline (prevalent cases).

### Air pollution exposure assessment

We used estimates of annual mean levels of exposure to air pollutants (PM_2.5_, BC, and NO_2_) from the 100 × 100 m European-wide hybrid land-use regression (LUR) models, which were developed within the ELAPSE project [19]. Briefly, the LUR models were developed based on monitoring measurements from the European Environment Agency AirBase for PM_2.5_ and NO_2_ in 2010 and from the ESCAPE project for BC. The annual mean PM_2.5_ absorbance data collected in 2009 and 2010, by filter reflectance, were considered as the annual mean concentration of BC for 2010. The models included satellite air pollution estimates, dispersion model estimates, land-use, road, and population density data as predictors. The spatial variations in the measured concentrations were explained by 66% for PM2.5, 51% for BC, and 58% for NO_2_. In the ELAPSE project, 2010 was selected as the year of air pollution assessment modeling because this was the earliest year providing sufficiently broad coverage of PM_2.5_ monitoring data across Europe. For consistency, 2010 was also used for NO_2_.

### Statistical analysis

The methodology applied in this study closely followed the protocol established by the ELAPSE project [20, 21]. Cox proportional hazard models, with age as the underlying timescale, were used to investigate the association between long-term exposure to air pollution and the incidence of asthma, COPD, and OAD. The models considered air pollution exposure and covariates as time-invariant factors. The follow-up period began on January 1, 2000, and ended at the earliest of the following dates: the date of the event (asthma, COPD, or OAD incidence), death, emigration, or December 31, 2018. All analyses accounted for data clustering within the same parish. The proportional hazard assumption for covariates and pollutants in the main models was assessed using a statistical test based on the scaled Schoenfeld residuals and their shapes against time.

Models were sequentially adjusted for pre-selected confounders. Model 1 included age (time scale) and sex (strata); Model 2 further adjusted for individual demographic and SES factors, including household income (categorized into deciles), employment status [unemployed, employed, or having support or pension (sick or cash support, under education, pension, others)], origin (Danish, immigrants/descendants from Western, or those from non-Western countries), marital status (unmarried, divorced, widowed, or married/registered partnership), and the highest educational level (primary, upper secondary, vocation/qualifying, vocational bachelor/short-cycle higher education, or more than college level); Model 3 included additional adjustments for regional average household income and unemployment rate, and the difference in these metrics between parish and region; Model 4, as our main model, further indirectly adjusting for health behavior-related factors (i.e., smoking status and BMI) not captured in the nationwide administrative cohort [22] by mathematically recalibrating hazard ratios (HRs) from Model 3 using the data from the Danish National Health Survey (DNHS) [23] in 2010 and 2013 (details in Text S1 in Supplementary materials). To examine the shape of the association, a natural spline term for each pollutant with two degrees of freedom was included in Model 4, replacing the linear term; the parish clustering term was excluded due to computational time. Two pollutant models were applied for all pollutant-pairs except for NO_2_ and BC due to a very high correlation (Spearman’s correlation; ρ=0.89). As a sensitivity analysis, we fit models with extrapolated time-varying and baseline year exposure (details in e 2 in Supplementary materials).

The effects of air pollution exposure were presented as HR with 95% confidence interval (CI) per interquartile range (IQR) increase of each pollutant. All statistical analyses and graphical presentations were conducted using R version 4.3.1 (R Project for Statistical Computing).

## RESULTS

From 3,323,612 individuals, we excluded 31,593 due to missing information on covariates, and 9,498 with pre-existing all three conditions (asthma, COPD, and OAD medication) at baseline, resulting in a base cohort of 3,282,521. From this, we defined three analytic sub-cohorts 3,248,586 for asthma, 3,240,669 for COPD, and 2,980,879 for OAD medication, after excluding individuals with prior events for each respective outcome (details in Figure S1 in Supplementary materials).

During a mean follow-up of 15.6 years, 55,637 individuals developed asthma, and 157,452 individuals developed COPD. During a mean follow-up of 14.9 years, 393,211 individuals were prescribed OAD medication (Table 1). Women were more likely to develop asthma or to be prescribed OAD medication, with the most pronounced sex difference seen for asthma (Table 1). While asthma incidence did not vary substantially by income, education, or employment status, COPD incidence were higher in the lowest SES groups. Asthma incidence was proportionally higher among immigrants/descendants from non-Western countries, while COPD was more common among those of Danish origin. COPD cases were more likely to be divorced or widowed, while smaller differences by marital status were observed for asthma patients. Individuals who developed asthma, COPD, or received OAD medication generally had slightly higher exposures compared to those who did not develop these outcomes. Baseline pollutant levels were relatively low: mean 12.4 µg/m^3^ for PM_2.5_, 20.2 µg/m^3^ for NO_2,_ and 1.0×10^-5^/m for BC (Table 2), but with mean values of PM_2.5_ and NO_2_ above WHO Guidelines [2].

**Table 1.**
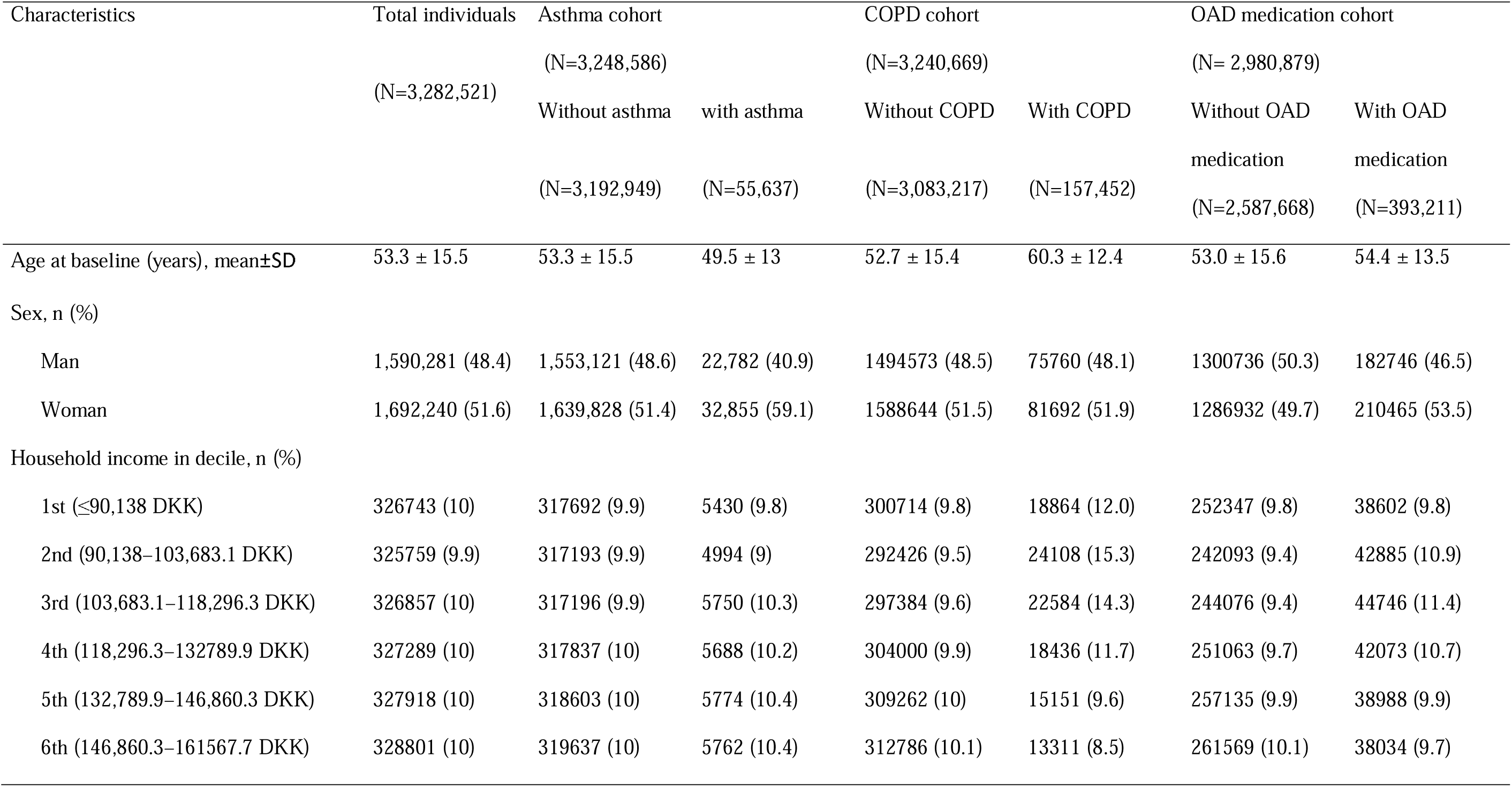
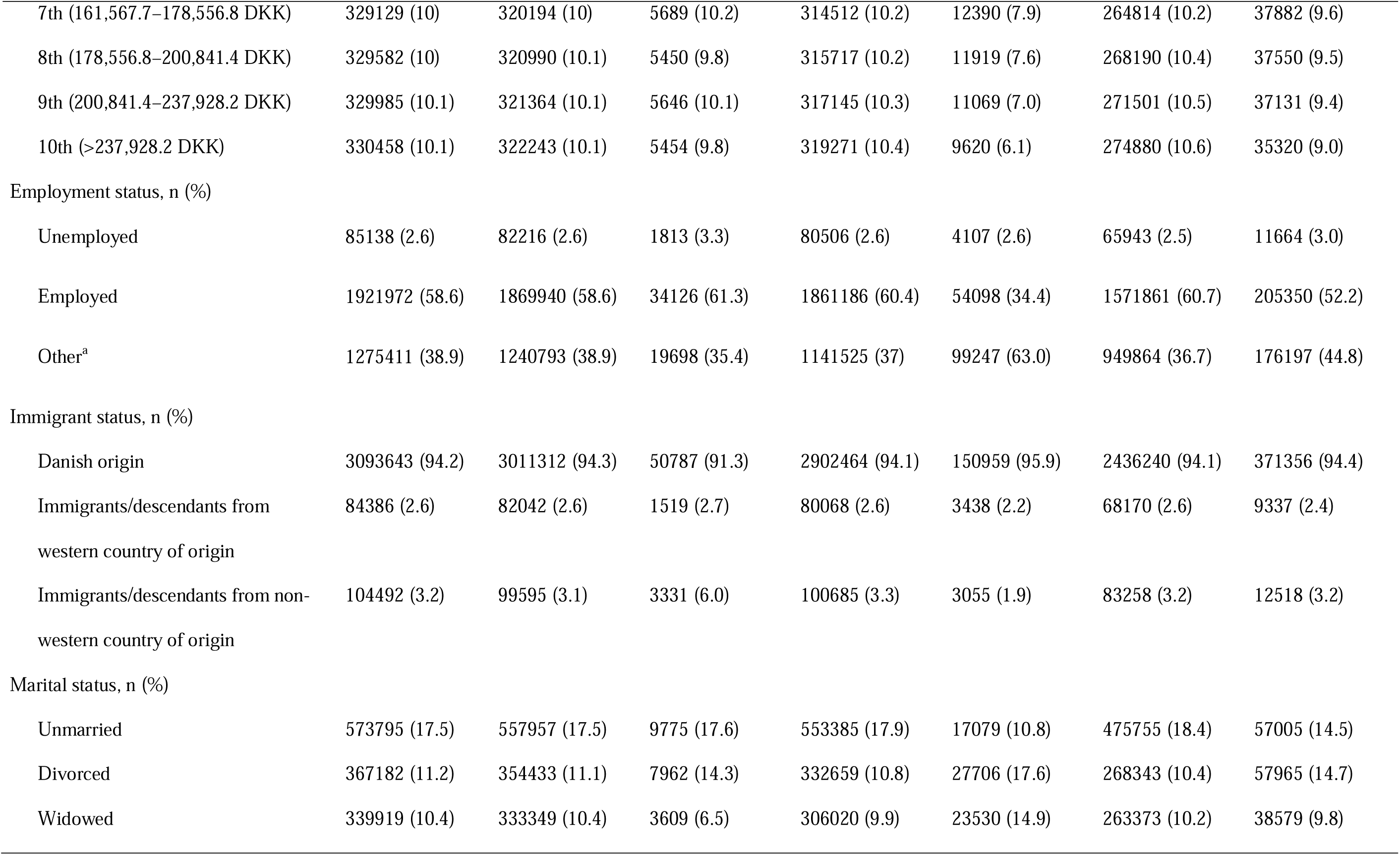
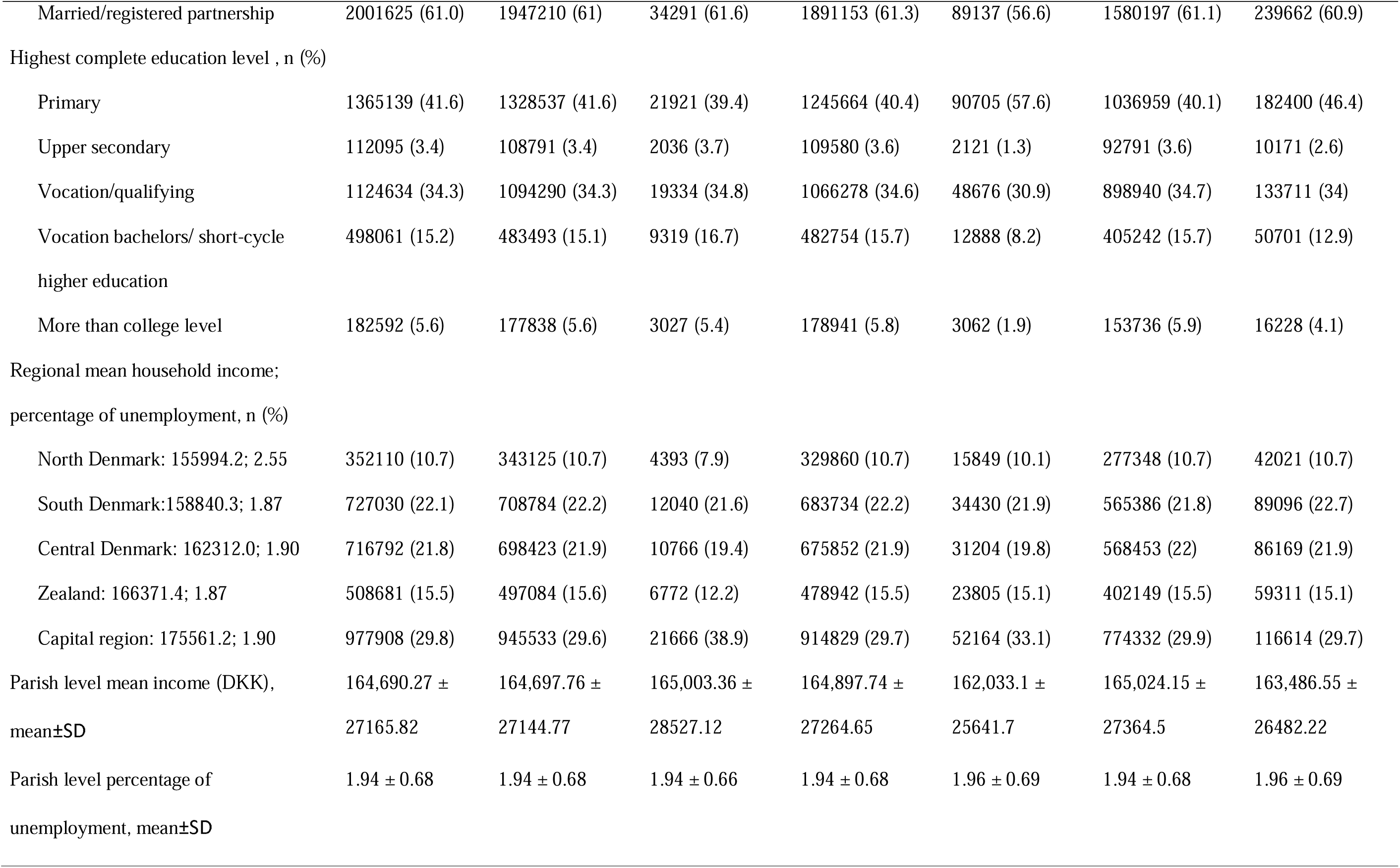
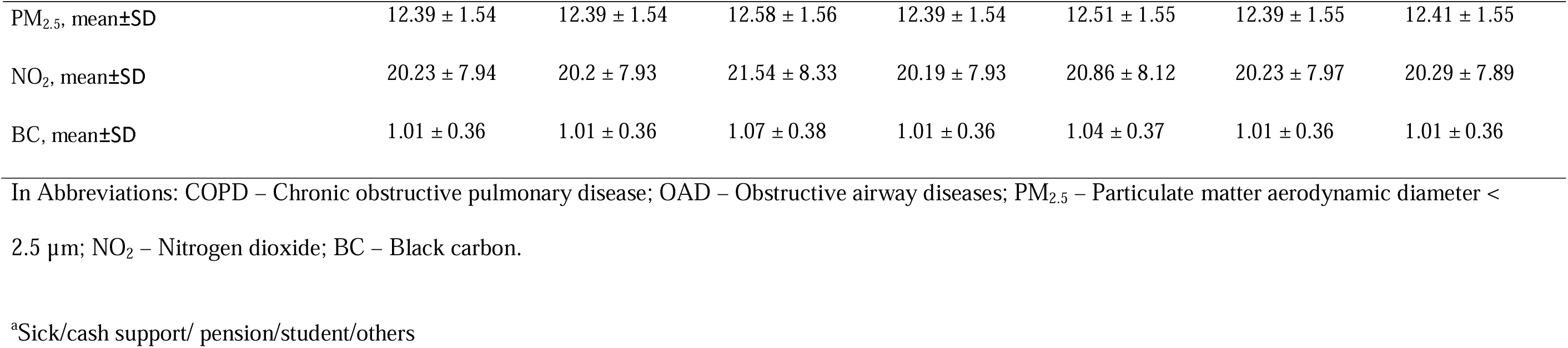
Demographic and socioeconomic characteristics of 3,282,521 individuals from the Danish nationwide administrative cohort at baseline (2000) by the status (having asthma, COPD, or OAD medication) at the end of follow-up.

**Table 2.** Descriptive characteristics for levels of annual mean exposure to ambient air pollution at baseline for total population (n=3,282,521)

| Exposure | Mean ± SD | IQR | Min | Percentile | | | | | Max | Spearson's rank correlation coefficients<br>($\rho$) | | |
| --- | --- | --- | --- | --- | --- | --- | --- | --- | --- | --- | --- | --- |
|  |  |  |  | 5<sup>th</sup> | 25<sup>th</sup> | 50<sup>th</sup> | 75<sup>th</sup> | 95<sup>th</sup> |  | PM<sub>2.5</sub> | NO<sub>2</sub> | BC |
| PM<sub>2.5</sub>, $\mu\text{g}/\text{m}^3$ | 12.4 ± 1.54 | 2.0 | 5.8 | 10 | 11.3 | 12.3 | 13.3 | 15.3 | 19.6 | 1.00 | - | - |
| NO<sub>2</sub>, $\mu\text{g}/\text{m}^3$ | 20.2 ± 7.94 | 10.3 | 3.7 | 9.4 | 14.5 | 18.6 | 24.8 | 35.7 | 72.2 | 0.60 | 1.00 | - |
| BC, 10<sup>-5</sup>/m | 1.0 ± 0.36 | 0.5 | 0.1 | 0.5 | 0.7 | 0.9 | 1.2 | 1.7 | 3.7 | 0.71 | 0.89 | 1.00 |

In Abbreviations: SD – standard deviation; IQR- Interquartile range; PM<sub>2.5</sub> – Particulate matter aerodynamic diameter < 2.5 $\mu\text{m}$ ; NO<sub>2</sub> – Nitrogen dioxide; BC
– Black carbon.
The minimum and maximum values are averages of the lowest and highest five values, respectively, to comply with General Data Protection Regulation.

We found statistically significant positive associations of PM_2.5_, BC, and NO_2_ with all three outcomes (Table 3), strongest for asthma and weakest for OAD medication. HRs attenuated from Model 1 to Model 4, by around half for asthma and OAD (not for NO_2_), and by around two-thirds for COPD, for which indirect adjustment for smoking and BMI had the biggest impact on the HR. We observed the strongest associations for asthma and COPD incidence with BC, which remained after adjustment for PM_2.5_ (Table 4). Associations with PM_2.5_ were weaker than those with BC and attenuated (asthma) or diminished after adjustment for BC (for COPD and OAD) (Table 4). Associations with NO_2_ were similar to those with BC, and the associations were independent from PM_2.5_ in two pollutant models. All associations were linear, with some flattening of the curve at higher levels (Figure 1). Sensitivity analyses with time-varying and base line year exposures presented robust results (Table S1 in Supplementary materials).

**Figure 1.**
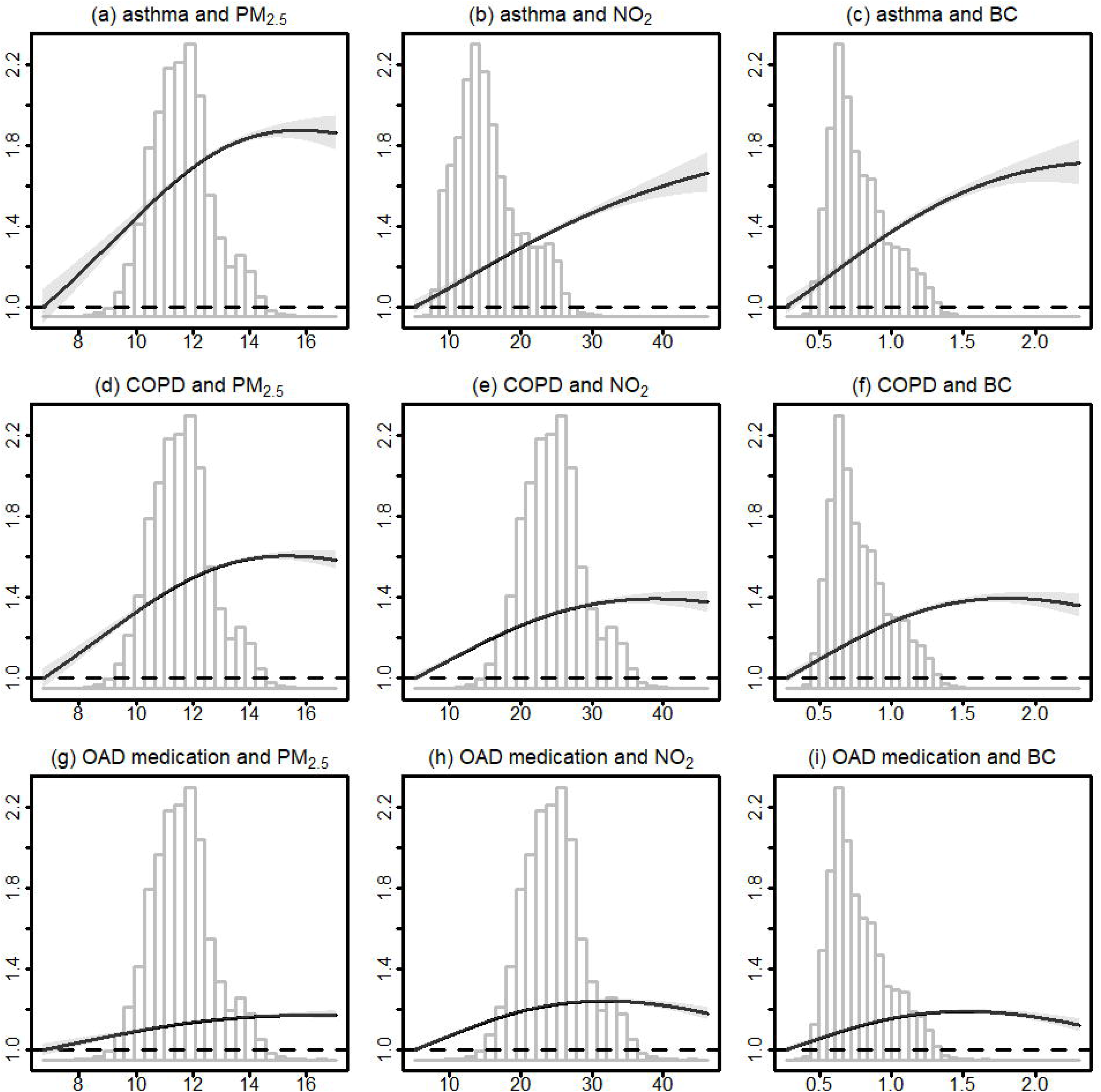
Exposure-response curve for the association between long-term exposure to air pollution and incidence of asthma, chronic obstructive pulmonary disease, and usage of medication for obstructive airway disease in the Danish administrative cohort study. Abbreviations: PM_2·5_ – Particulate matter aerodynamic diameter < 2·5 µm; NO_2_ – Nitrogen dioxide; BC – Black carbon. Note: Natural cubic splines with two degrees of freedom were fit for air pollutants to evaluate the shape of the associations based on the main model (Model 4) considering sex in a strata term and adjusted for household income, occupational status, immigrant status, marital status, and highest completed education level, regional mean household income, regional percentage of unemployment, and the difference of mean household income and percentage of unemployment, between parish and region, and indirectly adjusted for smoking status and body mass index. Solid lines indicate hazard ratio values, and gray shaded areas indicate their 95% confidence intervals. Dashed horizontal lines are the HRs equal to 1. Histograms with gray lines indicate the distribution of air pollutants. We exclude the extreme 0.1% values (i.e., the lowest 0.05% and the highest 0.05%) based on air pollutants’ values in the figure presentation.

**Table 3.**
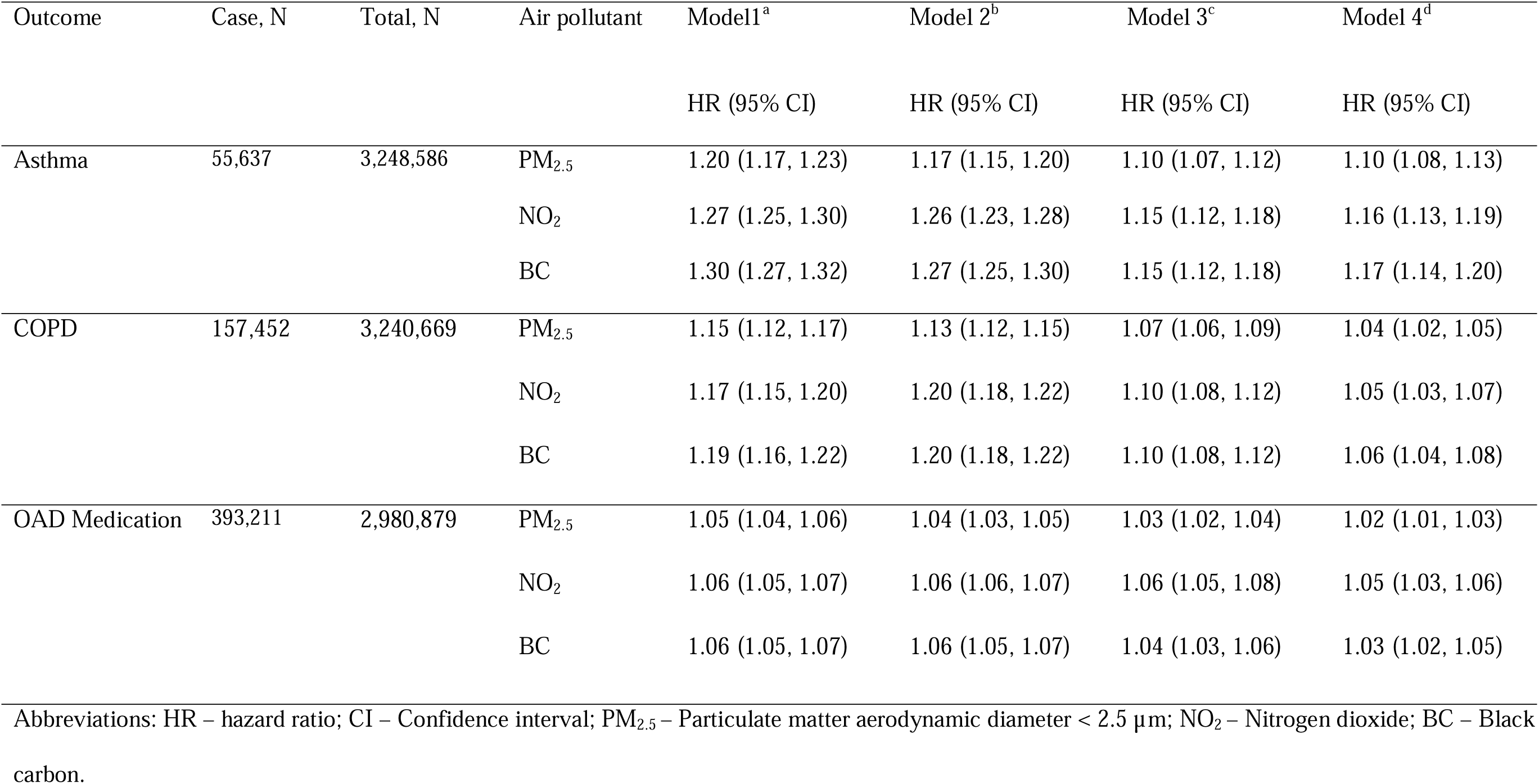
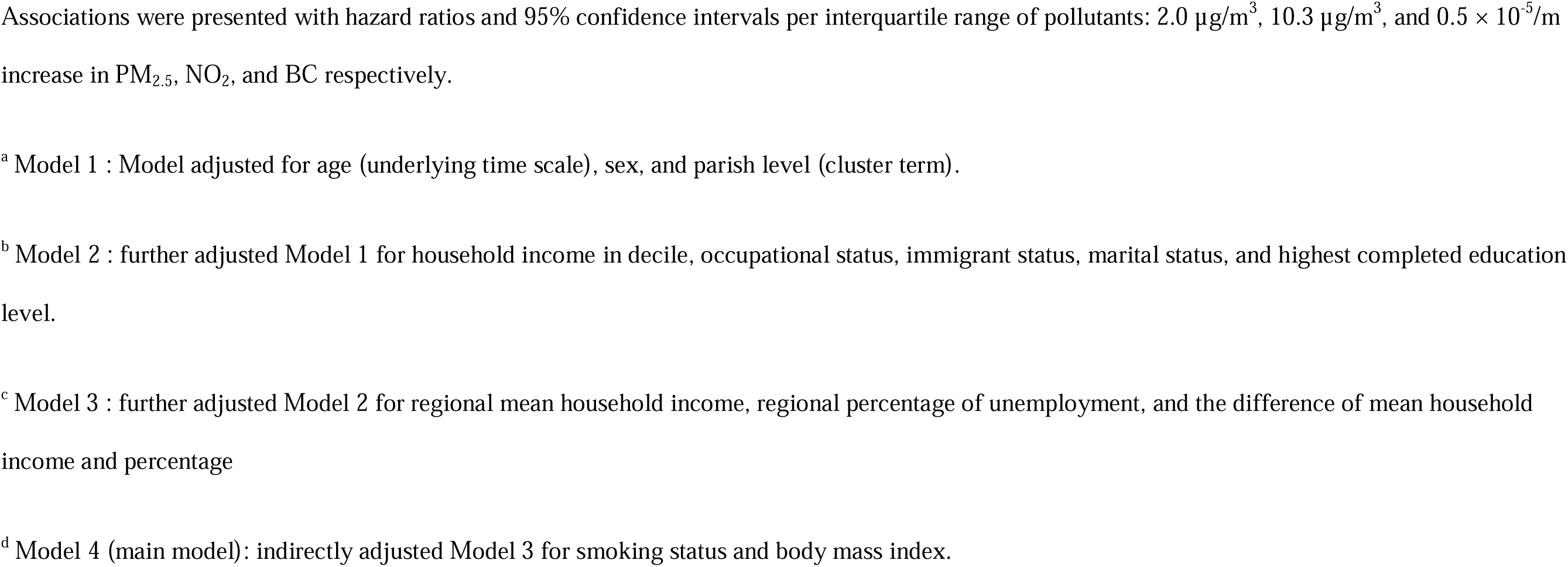
Associations between air pollution and incidence of asthma or chronic obstructive pulmonary disease, and usage of medication for obstructive airway disease in the Danish administrative cohort study.

| Outcome | Case, N | Total, N | Air pollutant | Model 1<sup>a</sup> | Model 2<sup>b</sup> | Model 3<sup>c</sup> | Model 4<sup>d</sup> |
| --- | --- | --- | --- | --- | --- | --- | --- |
|  |  |  |  | HR (95% CI) | HR (95% CI) | HR (95% CI) | HR (95% CI) |
| Asthma | 55,637 | 3,248,586 | PM<sub>2.5</sub> | 1.20 (1.17, 1.23) | 1.17 (1.15, 1.20) | 1.10 (1.07, 1.12) | 1.10 (1.08, 1.13) |
|  |  |  | NO<sub>2</sub> | 1.27 (1.25, 1.30) | 1.26 (1.23, 1.28) | 1.15 (1.12, 1.18) | 1.16 (1.13, 1.19) |
|  |  |  | BC | 1.30 (1.27, 1.32) | 1.27 (1.25, 1.30) | 1.15 (1.12, 1.18) | 1.17 (1.14, 1.20) |
| COPD | 157,452 | 3,240,669 | PM<sub>2.5</sub> | 1.15 (1.12, 1.17) | 1.13 (1.12, 1.15) | 1.07 (1.06, 1.09) | 1.04 (1.02, 1.05) |
|  |  |  | NO<sub>2</sub> | 1.17 (1.15, 1.20) | 1.20 (1.18, 1.22) | 1.10 (1.08, 1.12) | 1.05 (1.03, 1.07) |
|  |  |  | BC | 1.19 (1.16, 1.22) | 1.20 (1.18, 1.22) | 1.10 (1.08, 1.12) | 1.06 (1.04, 1.08) |
| OAD Medication | 393,211 | 2,980,879 | PM<sub>2.5</sub> | 1.05 (1.04, 1.06) | 1.04 (1.03, 1.05) | 1.03 (1.02, 1.04) | 1.02 (1.01, 1.03) |
|  |  |  | NO<sub>2</sub> | 1.06 (1.05, 1.07) | 1.06 (1.06, 1.07) | 1.06 (1.05, 1.08) | 1.05 (1.03, 1.06) |
|  |  |  | BC | 1.06 (1.05, 1.07) | 1.06 (1.05, 1.07) | 1.04 (1.03, 1.06) | 1.03 (1.02, 1.05) |

Abbreviations: HR – hazard ratio; CI – Confidence interval; PM<sub>2.5</sub> – Particulate matter aerodynamic diameter < 2.5 µm; NO<sub>2</sub> – Nitrogen dioxide; BC – Black carbon.
Associations were presented with hazard ratios and 95% confidence intervals per interquartile range of pollutants: $2.0 \mu\text{g}/\text{m}^3$ , $10.3 \mu\text{g}/\text{m}^3$ , and $0.5 \times 10^{-5}/\text{m}$ increase in $\text{PM}_{2.5}$ , $\text{NO}_2$ , and BC respectively.
<sup>a</sup> Model 1 : Model adjusted for age (underlying time scale), sex, and parish level (cluster term).
<sup>b</sup> Model 2 : further adjusted Model 1 for household income in decile, occupational status, immigrant status, marital status, and highest completed education level.
<sup>c</sup> Model 3 : further adjusted Model 2 for regional mean household income, regional percentage of unemployment, and the difference of mean household income and percentage
<sup>d</sup> Model 4 (main model): indirectly adjusted Model 3 for smoking status and body mass index.

**Table 4.**
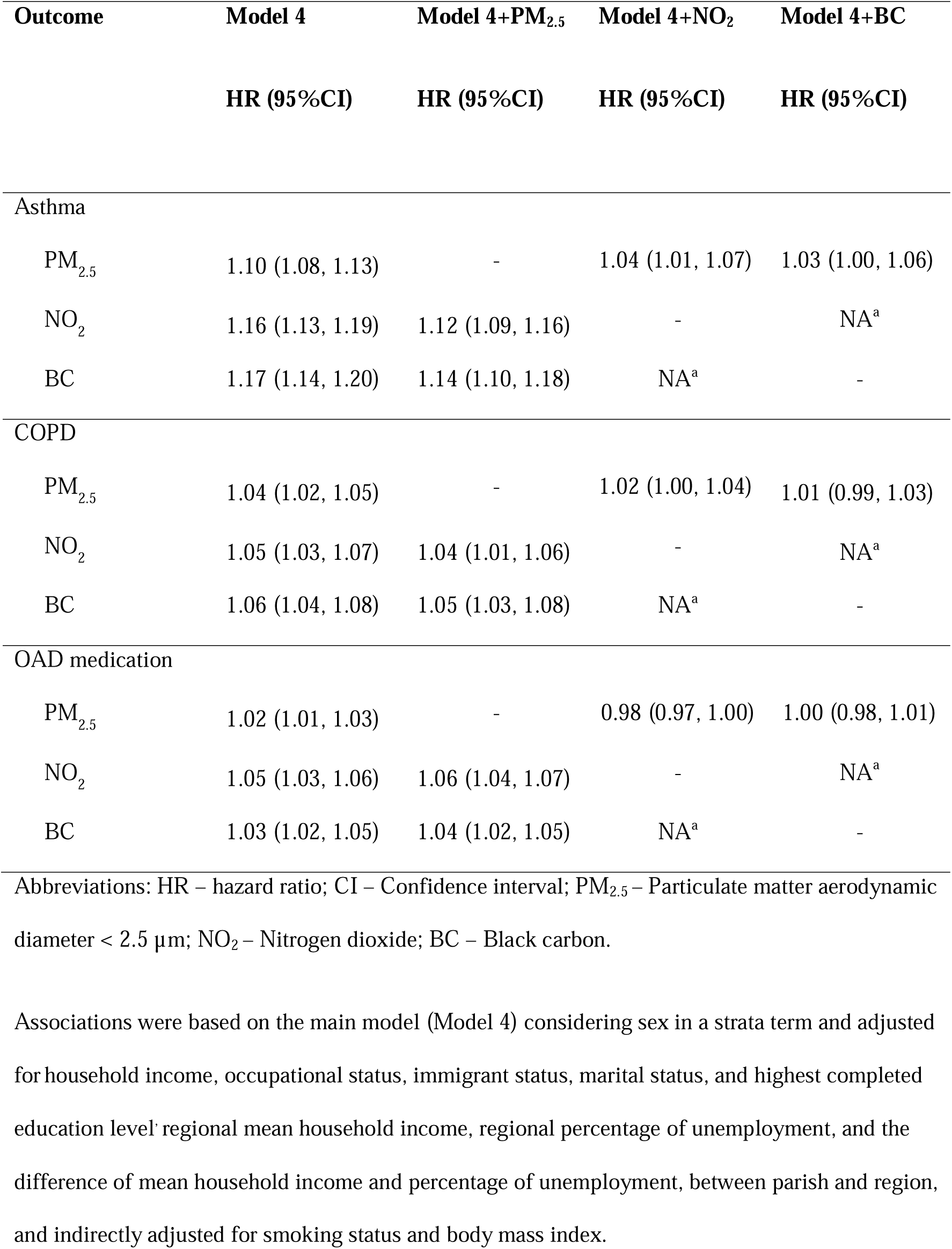
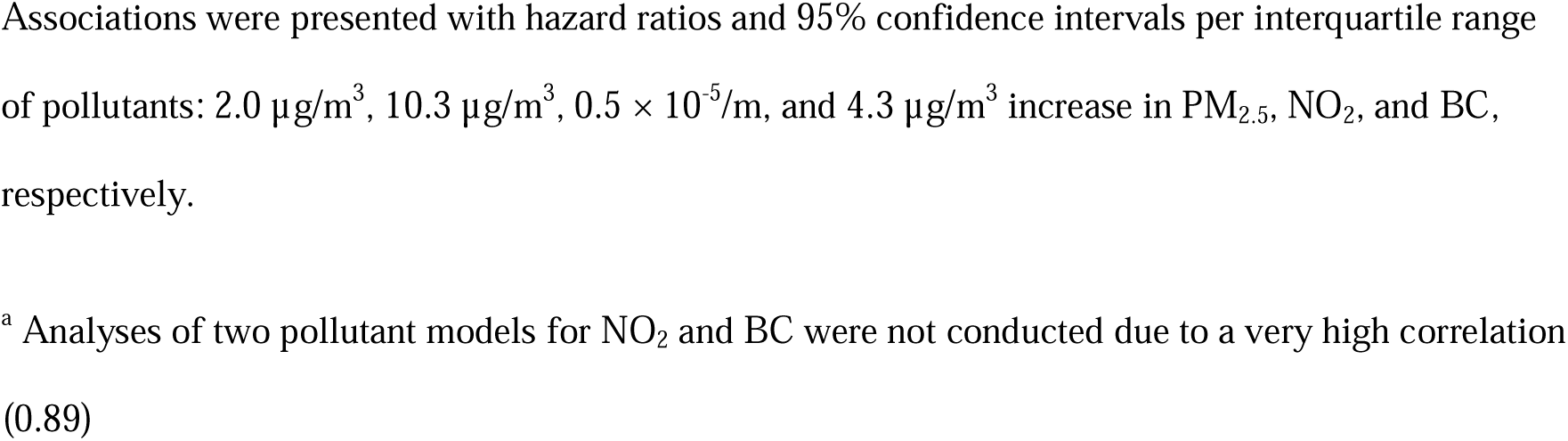
Associations between long-term exposure to air pollution and incidence of asthma, chronic obstructive pulmonary disease, and usage of medication for obstructive airway disease in the Danish administrative cohort study.

| Outcome | Model 4 | Model 4+PM<sub>2.5</sub> | Model 4+NO<sub>2</sub> | Model 4+BC |
| --- | --- | --- | --- | --- |
|  | HR (95%CI) | HR (95%CI) | HR (95%CI) | HR (95%CI) |
| Asthma |  |  |  |  |
| PM<sub>2.5</sub> | 1.10 (1.08, 1.13) | - | 1.04 (1.01, 1.07) | 1.03 (1.00, 1.06) |
| NO<sub>2</sub> | 1.16 (1.13, 1.19) | 1.12 (1.09, 1.16) | - | NA<sup>a</sup> |
| BC | 1.17 (1.14, 1.20) | 1.14 (1.10, 1.18) | NA<sup>a</sup> | - |
| COPD |  |  |  |  |
| PM<sub>2.5</sub> | 1.04 (1.02, 1.05) | - | 1.02 (1.00, 1.04) | 1.01 (0.99, 1.03) |
| NO<sub>2</sub> | 1.05 (1.03, 1.07) | 1.04 (1.01, 1.06) | - | NA<sup>a</sup> |
| BC | 1.06 (1.04, 1.08) | 1.05 (1.03, 1.08) | NA<sup>a</sup> | - |
| OAD medication |  |  |  |  |
| PM<sub>2.5</sub> | 1.02 (1.01, 1.03) | - | 0.98 (0.97, 1.00) | 1.00 (0.98, 1.01) |
| NO<sub>2</sub> | 1.05 (1.03, 1.06) | 1.06 (1.04, 1.07) | - | NA<sup>a</sup> |
| BC | 1.03 (1.02, 1.05) | 1.04 (1.02, 1.05) | NA<sup>a</sup> | - |

Abbreviations: HR – hazard ratio; CI – Confidence interval; PM<sub>2.5</sub> – Particulate matter aerodynamic diameter < 2.5 µm; NO<sub>2</sub> – Nitrogen dioxide; BC – Black carbon.
Associations were presented with hazard ratios and 95% confidence intervals per interquartile range of pollutants: 2.0 $\mu\text{g}/\text{m}^3$ , 10.3 $\mu\text{g}/\text{m}^3$ , $0.5 \times 10^{-5}/\text{m}$ , and 4.3 $\mu\text{g}/\text{m}^3$ increase in $\text{PM}_{2.5}$ , $\text{NO}_2$ , and BC, respectively.
<sup>a</sup> Analyses of two pollutant models for $\text{NO}_2$ and BC were not conducted due to a very high correlation (0.89)

## DISCUSSION

In a large Danish nationwide analysis conduted in low-exposure setting, we found that long-term exposure to PM_2.5_, BC, and NO_2_ were associated with the incidence of asthma and COPD, with the strongest associations observed for asthma. We present novel findings of the assocation of BC with asthma and COPD, stronger than and independent of those with PM_2.5_.

Our findings for BC and COPD align with four earlier studies all showing positive associations[12–15], including one from three ELAPSE cohorts showing that the association persisted after adjustment for PM_2.5_ [12]. Our results on BC and asthma are in agreement with the ELAPSE study, which used three cohorts and employed an identical asthma incidence definition, and found similar HRs as in our study [HR: 1.15 (95% CI, 1.08-1.23) per 0.5×10^-5^/m] [10]. Liu et al. also found that BC remained robust (even became stronger) in the two pollutant model with PM_2.5_, whereas the PM_2.5_ estimate diminished upon BC adjustment [10]. In contrast, the ESCAPE study, based on six cohorts and relying on self-reported asthma, found no statistically significant association with BC [odds ratio: 1.03 (95% CI, 0.97-1.09) per 0.5×10^-5^/m [11]. This estimate is comparable to our finding for BC and OAD medication (HR of 1.03, Table 3), but much weaker than our finding for asthma incidence defined as first hospital contact (HR: 1.17). This indicates that air pollution either is linked to more clinically confirmed asthma, captured by hospital contact than by medication use or self-reports, which leads to overestimated incidence [24, 25], including milder or misdiagnosed cases. We see a similar trend for COPD, with stronger associations with COPD defined as hospital than by OAD prescriptions.

Our findings show the most robust associations with BC and NO_2_, suggesting the relevance of locally generated combustion-related sources - particularly from biomass burning and traffic - with adverse respiratory outcomes. Current understanding suggests that NO_2_, an airway irritant linked to airway inflammation and airflow limitation [26], may act both as a causal agent in asthma/COPD development and as a proxy for traffic-related PM_2.5_, BC or ultrafine particles (UFPs), which can deposit in the respiratory tract and the lung alveoli and include oxidative stress, inflammation, and other biochemical changes related to asthma/COPD [27]. A recent review described mechanistic pathways of how PM, BC, and gases can disrupt the airway epithelial barrier, exacerbating inflammatory responses and airway remodeling; key features in pathology of asthma and COPD [28]. Due to the high correlation, we could not distinguish between BC and NO_2_ effects in two pollutant models and assume independent effects of both pollutants. Our finding on the effect of PM_2.5_ and NO_2_ on asthma and COPD, independently of each other, were also reported in another large administrative Canadian study [29].

The main strength of our study is the large and unselected cohort of 3.3 million Danish adults followed over 18 years, with high-resolution nationwide air pollution data (100×100 m), validated definitions of asthma and COPD incidence and detailed data on SES, from unique Danish nationwide hospital, medication, education, and employment registers, allowing for the first nationwide study of air pollution and incidence of asthma and COPD in Denmark. While Asthma and COPD incidence defined based on hospital discharge diagnoses may capture more clinically confirmed cases, they may also underestimate the true incidence, as not all patients require a hospital contact. However, this definition is appealing as it is typically confirmed by objective measurements of lung function and reversible airflow obstruction, as standard procedures in Danish hospitals. The high specificity of asthma (0.98) [30] and COPD (0.92) [31] diagnoses in the Danish National Patient Registry further supports their validity for use in epidemiological studies.

Our study has several limitations. We lacked information on air pollution during any outdoor activity, at the workplace, or related to commuting. Our exposure assessment was based on 2010 LUR models applied to participants’ home addresses in 2000, which may introduce exposure misclassification, particularly if pollutant levels changed over time and participants moved during follow-up. This approach was due to the limited monitoring data in earlier years to develop models for PM_2·5_.

However, a previous study found [19] showed that 2010 model predictions were strongly correlated with those from 2000 for NOL (R² > 0.84) and moderately for PML.L with 2013 data (R² = 0.49), indicating relatively stable spatial contrasts over time and supporting the use of 2010 exposure estimates in our analysis. Additionally, sensitivity analyses accounting for temporal trends and residential mobility yielded consistent results (Tables S1).

Furthermore, as in many studies using administrative databases, we lacked information on individual-level lifestyle risk factors. To address this, we applied indirect adjustments for smoking status and BMI, showing an expected impact on the HRs, as seen in studies with individual-level smoking, BMI, etc. [10, 12, 14, 15]. Furthermore, it is still important to note that the validity of the indirect adjustment method depends on how well the ancillary dataset (DNHS 2010) represents our cohort: supporting this, the 2010 DNHS showed a similar distribution of demographic and SES variables as the Danish nationwide administrative cohort (Table S2 in Supplementary materials), except for higher education and income in survey participants.

## CONCLUSION

In a large Danish nationwide analysis of more than 3 million adults living in a low-exposure setting, we showed that long-term exposure to PM_2.5_, BC, and NO_2_ is associated with the development of asthma and COPD, with the strongest associations with asthma. We presented novel findings of a strong association of BC with asthma and COPD incidence, independent of that of PM_2.5_. Our findings emphasise that air pollution reduction should be a priority for chronic lung disease prevention strategies and that regulation of BC may further help reduce the burden of lung disease.

## Data Availability

The authors do not have permission to share data.

## Acknowledgements

The presented work was funded by Clean Air Fund. Rina So received a grant LUNDBECK foundation (#R449-2023-1412). The ELAPSE was supported by funding from HEI Research Agreement (#4954-RFA14-3/16-5-3), jointly funded by the United States Environmental Protection Agency (EPA) (Assistance Award No. R82811201). They were not involved in the analysis, interpretation or drafting of the manuscript.

## Author contributions

The study was conceptualised and designed by Z.J. Andersen, R. So, G. Hoek, B. Brunekreef, G. Hoek and B. Brunekreef are Principal Investigators of the ELAPSE project. Statistical analysis and drafting of the manuscript was conducted by R So, Z.J. Andersen helped R So. in drafting the manuscript, and J. Zhang helped in summarizing current evidence. R So. prepared the individual cohort data for the analyses. C.S. Ulrik helped with the definition of the asthma and COPD and clinical interpretation of the findings. G. Hoek, B. Brunekreef, and J. Chen coordinated the ELAPSE project, M. Staffoggia, J. Chen and E. Samoli contributed to the statistical analysis strategy and scripts for the statistical analyses. K. de Hoogh and J. Chen worked for the exposure assessment. All authors have read and revised the manuscript for important intellectual content, and contributed to the interpretation of the results. All authors have approved the final draft of the manuscript.

