## Supplementary for "Long-term exposure to air pollution and risk of adult-onset asthma and COPD: Danish nationwide cohort study"

**Figure S1. Flowchart of study population**

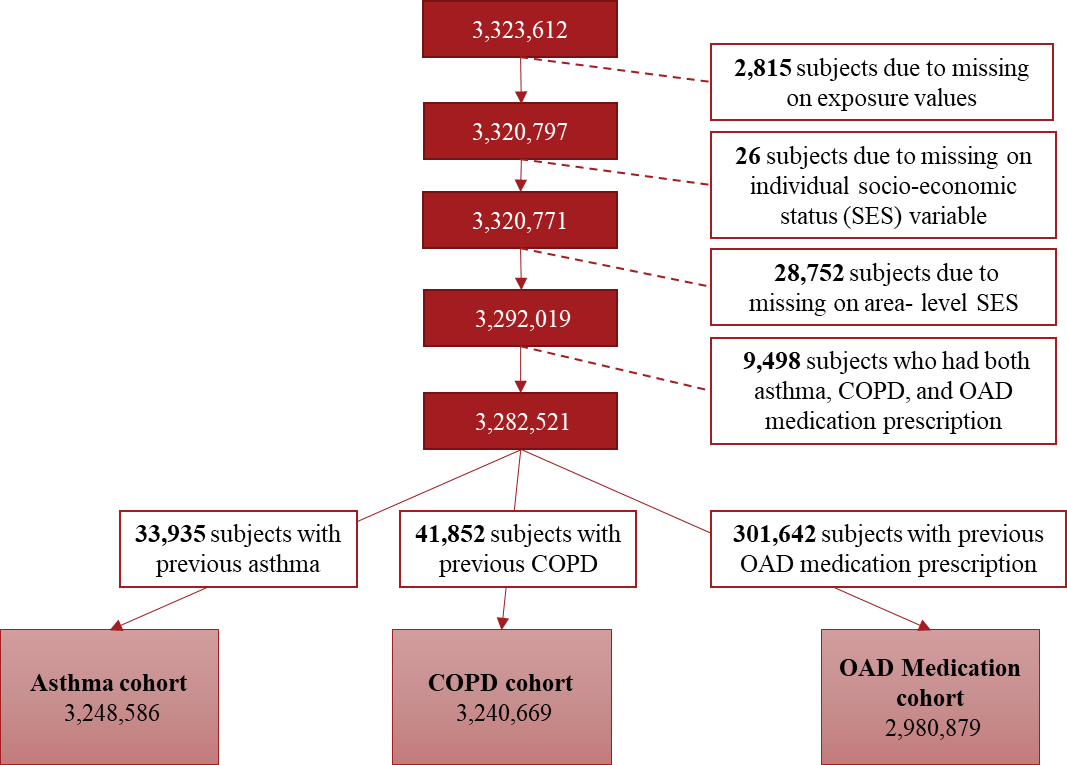

Abbreviation: **SES** - Socioeconomic Status; **COPD** - Chronic Obstructive Pulmonary Disease; **OAD** -Obstructive Airway Disease

Note: **Dark red boxes** represent the total number of individuals at each step of the selection process. **White boxes** indicate exclusion criteria and the number of excluded subjects. The **light red boxes** at the bottom show the final sample size for each analytic cohort.

**Text S1. Indirect adjustment**

We implemented the indirect adjustment method proposed by Shin et al.^1^, which uses ancillary survey data representative of the cohort to adjust hazard ratio (HR) estimates for missing confounders in the administrative cohort. This method requires two pieces of information: (1) the relationship between air pollution exposure and lifestyle factors from an external population; (2) a risk function for these lifestyle factors in relation to the outcome of interest. These should come from independent sources.^1^ Therefore, we derived these information from the Danish National Health Survey (DNS)^2^ in 2010 and 2013, separately, each based on different random samples of the population.

The indirect adjustment method was performed with the following steps.

- First, as suggested in Shin et al., we compared the distributions of air pollution, age, sex, and other variables between the administrative cohort and the survey (Table S2), to evaluate the representativeness of ancillary survey data (the DNS in 2010 in our study) to the cohort study.
- Next, to get (1) mentioned above, we examined whether smoking status and body mass index (BMI) were associated with air pollution estimates using multiple linear regression while adjusting for other covariates in Model 3 in the DNS in 2010 (Table S3). For (2) mentioned above, we then obtained the HR estimates of smoking status and BMI for an outcome of interest from the Cox model, adjusting for other potential covariates in the DNS 2013 (Table S4).
- Lastly, the adjusted parameter is calculated as $\tilde{\beta}=\hat{\gamma}-\bar{\Delta}\tilde{\lambda}$, where $\hat{\gamma}$is the unadjusted effect estimate from Model 3, $\bar{\Delta}$ is the matrix of associations between observed variables and missing covariates in the administrative cohort, obtained from the DNS in 2010, and $\tilde{\lambda}$ is the vector of risk estimates of smoking status and BMI on health outcome derived from the DNS in 2013. The variance of the adjusted estimates accounts for the variance of the observed variables, the variance-covariance matrix of the association between missing factors and health outcome from the DNS in 2013, and the variance-covariance matrix of associations between observed variables and missing factors from the DNS in 2010.

**Text S2. Backward and forward extrapolation method for time-varying exposure estimates.**

As part of a sensitivity analysis, annual mean levels of PM_2.5_, NO_2_, and BC for each year during follow-up were estimated using a backward- and forward-extrapolation method, based on estimates from the Danish Eulerian Hemispheric Model (DEHM)^3^. A detailed description of the method can be found elsewhere^4,5^. In brief, DEHM offers monthly mean concentration estimates at a spatial resolution of 26 km × 26 km across Europe, back to at least 1990. DEHM was utilized due to the lack of continuous monitoring data from AirBase during the study period, with both datasets showing consistent temporal trends. Within the ELAPSE project, population-weighted annual mean air pollution concentrations were calculated at the NUTS-1 level (all of Denmark is within one NUTS-1 region). These values were then used to calculate the ratios of exposure levels for each follow-up year relative to 2010, which were subsequently multiplied by the LUR model exposure estimates at participants’ residential addresses at each year.

**Table S1. The comparison of hazard ratios associated with long-term exposure to air pollutants between models with exposure estimates in 2010 at baseline addresses and those with back-extrapolated time-varying exposure estimates in the Danish administrative cohort.**

| **Population** | **Individuals, N** | **Cases, N** | **Exposure estimation type** | **PM_2.5_ HR (95% CI)** | **NO_2_ HR (95% CI)** | **BC HR (95% CI)** |
| --- | --- | --- | --- | --- | --- | --- |
| Asthma dataset | 3,248,586 | 55,637 | Exposure in 2010 (main) | 1.10 (1.08, 1.13) | 1.16 (1.13, 1.19) | 1.17 (1.14, 1.20) |
|  |  |  | Time-varying exposure | 1.09 (1.08, 1.10) | 1.18 (1.16, 1.20) | 1.21 (1.19, 1.23) |
|  |  |  | Extrapolated to baseline | 1.06 (1.05, 1.08) | 1.11 (1.09, 1.13) | 1.16 (1.13, 1.19) |
| COPD dataset | 3,240,669 | 157,452 | Exposure in 2010 (main) | 1.04 (1.02, 1.05) | 1.05 (1.03, 1.07) | 1.06 (1.04, 1.08) |
|  |  |  | Time-varying exposure | 1.04 (1.03, 1.05) | 1.08 (1.07, 1.09) | 1.11 (1.10, 1.12) |
|  |  |  | Extrapolated to baseline | 1.05 (1.04, 1.06) | 1.08 (1.06, 1.09) | 1.10 (1.08, 1.12) |
| OAD medication dataset | 2,980,879 | 393,211 | Exposure in 2010 (main) | 1.02 (1.01, 1.03) | 1.05 (1.03, 1.06) | 1.03 (1.02, 1.05) |
|  |  |  | Time-varying exposure | 1.01 (1.01, 1.02) | 1.04 (1.04, 1.05) | 1.04 (1.03, 1.05) |
|  |  |  | Extrapolated to baseline | 1.02 (1.01, 1.02) | 1.05 (1.04, 1.06) | 1.05 (1.03, 1.06) |

Abbreviations: PM_2.5_ – Particulate matter aerodynamic diameter < 2.5 µm; NO_2_ – Nitrogen dioxide; BC – Black carbon.

Associations were presented with hazard ratios and 95% confidence intervals per interquartile range of pollutants obtained from exposure in 2010 (main): 2.0 µg/m^3^, 10.3 µg/m^3^, and 0.5 × 10^-5^/m increase in PM_2.5_, NO_2_, and BC, respectively.

Hazard ratios and confidence intervals were adjusted for age (underlying time scale), sex (strata), parish level (cluster term), household income in decile, occupational status, immigrant status, marital status, and highest completed education level, regional mean household income, regional percentage of unemployment, and the difference of mean household income and percentage of unemployment, between parish and region, and indirectly adjusted for smoking status and body mass index.

For the models with time-varying exposure, we additionally included the strata term for each 5 years.

**Table S2.** **Comparison of descriptive statistics for demographic characteristics between the Danish nationwide administrative cohort and the Danish health survey, who were aged 30 or above.**

| **Characteristics** | **Danish nationwide administrative cohort (%)** | **Danish Health Survey 2010 (%)** |
| --- | --- | --- |
| Age |  |  |
| 30-39 | 24.3 | 20.6 |
| 40-49 | 22.2 | 23.4 |
| 50-59 | 22.1 | 20.4 |
| 60-69 | 14.4 | 19.8 |
| 70-79 | 10.9 | 10.4 |
| 80-89 | 5.3 | 4.9 |
| ≥90 | 0.9 | 0.5 |
| Sex |  |  |
| Men | 48.4 | 49.1 |
| Women | 51.6 | 50.9 |
| Household income in quintile^a^ |  |  |
| 1^st^ | 19.9 | 7.3 |
| 2^nd^ | 20.0 | 15.5 |
| 3^rd^ | 20.0 | 16.1 |
| 4^th^ | 20.0 | 21.9 |
| 5^th^ | 20.1 | 39.2 |
| employment status |  |  |
| Unemployed | 2.6 | 1.3 |
| Having support or pension | 38.9 | 37.3 |
| Employed | 58.6 | 61.4 |
| Immigrant status |  |  |
| Danish origin | 94.2 | 91.5 |
| Western country of origin | 2.6 | 4.1 |
| Non-Western country of origin | 3.2 | 4.5 |
| Marital status |  |  |
| Unmarried | 17.5 | 17.6 |
| Divorced | 11.2 | 11.6 |
| Widowed | 10.4 | 8.5 |
| Married/registered partnership | 61.0 | 62.3 |
| Highest complete education level |  |  |
| Primary | 41.6 | 26.7 |
| Upper secondary | 3.4 | 4.1 |
| Vocation/qualifying | 34.3 | 38.0 |
| Vocation bachelors/ short-cycle higher education | 15.2 | 21.2 |
| College level and over | 5.6 | 9.9 |
| Region |  |  |
| North Denmark | 10.7 | 10.5 |
| South Denmark | 22.1 | 21.6 |
| Central Denmark | 21.8 | 22.0 |
| Zealand | 15.5 | 16.1 |
| Capital region | 29.8 | 29.8 |
| Parish level mean income in quintile^a^ |  |  |
| 1^st^ | 19.1 | 6.1 |
| 2^nd^ | 18.7 | 6.5 |
| 3^rd^ | 19.7 | 13.1 |
| 4^th^ | 21.1 | 26.6 |
| 5^th^ | 21.4 | 47.8 |
| Parish level percentage of unemployment in quintile^b^ |  |  |
| 1^st^ | 19.9 | 48.9 |
| 2^nd^ | 19.4 | 17.9 |
| 3^rd^ | 19.9 | 15.5 |
| 4^th^ | 19.8 | 9.3 |
| 5^th^ | 20.9 | 8.5 |

^a^Comparison of household income and area-level mean household income between two datasets, based on quintiles from the Danish nationwide administrative cohort, considering the difference in price indexes due to the time difference.

^b^ Based on quintiles from the Danish nationwide administrative cohort

**Table S3. Relationship between air pollutants and lifestyle risk factors (smoking status and body mass index), used for the indirect adjustment, in the Danish National Health Survey 2010 aged 30 years or older (N=51,388).**

| **Lifestyle factor** | **PM_2.5_** | | | **NO_2_** | | | **BC** | | |
| --- | --- | --- | --- | --- | --- | --- | --- | --- | --- |
|  | **Beta** | **SE** | **p-value** | **Beta** | **SE** | **p-value** | **Beta** | **SE** | **p-value** |
| Smoking |  |  |  |  |  |  |  |  |  |
| Never smoker | Ref. | Ref. | Ref. | Ref. | Ref. | Ref. | Ref. | Ref. | Ref. |
| Previous smoker | 0.0035 | 0.0009 | 0.0001 | 0.0012 | 0.0002 | <.0001 | 0.0172 | 0.0045 | 0.0002 |
| Current smoker | 0.0043 | 0.0008 | <.0001 | 0.0012 | 0.0002 | <.0001 | 0.0155 | 0.0042 | 0.0002 |
| Body mass index (BMI) |  |  |  |  |  |  |  |  |  |
| Underweight (BMI<18.5) | 0.0001 | 0.0002 | 0.6795 | 0.0002 | 0.0001 | 0.0111 | 0.0026 | 0.0013 | 0.0419 |
| Normal (BMI:18.5-25) | Ref. | Ref. | Ref. | Ref. | Ref. | Ref. | Ref. | Ref. | Ref. |
| Overweight (BMI: 25-30) | -0.0022 | 0.0009 | 0.0139 | -0.0012 | 0.0002 | <.0001 | -0.0236 | 0.0047 | <.0001 |
| Obese (BMI>30) | -0.0039 | 0.0007 | <.0001 | -0.0011 | 0.0002 | <.0001 | -0.0263 | 0.0035 | <.0001 |

Abbreviation: SE – Standard error; Ref. - Reference; PM_2.5_ – Particulate matter aerodynamic diameter < 2.5 µm; NO_2_ – Nitrogen dioxide; BC – Black carbon.

Associations were adjusted for age, sex, household income in decile, occupational status, immigrant status, marital status, highest completed education level, regional mean household income, regional percentage of unemployment, and the difference of mean household income and percentage of unemployment, between parish and region.

**Table S4. The risk estimates for lifestyle risk factors (smoking status and body mass index) on asthma, COPD, and OAD medication, used for the indirect adjustment, in the Danish National Health Survey 2013 aged 30 years or older (125,184).**

| **Lifestyle factor** | **Asthma**  **(N=754)**  **HR (95% CI)** | **COPD (N=1489)**  **HR (95% CI)** | **OAD medication**  **(n=4105)**  **HR (95% CI)** |
| --- | --- | --- | --- |
| Smoking status |  |  |  |
| Never smoker | Ref. | Ref. | Ref. |
| Previous smoker | 1.21 (1.03, 1.41) | 4.96 (4.13, 5.97) | 1.81 (1.67, 1.95) |
| Current smoker | 0.83 (0.66, 1.03) | 9.44 (7.82, 11.40) | 3.02 (2.77, 3.28) |
| Body mass index (BMI) |  |  |  |
| Underweight (BMI<18.5) | 0.29 (0.09, 0.91) | 2.29 (1.79, 2.94) | 1.58 (1.29, 1.95) |
| Normal (BMI:18.5-25) | Ref. | Ref. | Ref. |
| Overweight (BMI: 25-30) | 1.18 (1.00, 1.40) | 0.85 (0.75, 0.95) | 1.05 (0.98, 1.13) |
| Obese (BMI>30) | 1.80 (1.49, 2.18) | 1.13 (0.98, 1.30) | 1.38 (1.27, 1.51) |

Abbreviation: HR- Hazard ratio; CI – Confidence interval; Ref. - Reference; PM2.5 – Particulate matter aerodynamic diameter < 2.5 µm; NO2 – Nitrogen dioxide; BC – Black carbon; COPD – chronic obstructive pulmonary disease; OAD – obstructive airway disease.

The risk estimates were obtained from Cox proportional hazard models including age (underlying time scale), sex (strata), smoking status, body mass index, household income in quintile, employment status, immigrant status, highest completed education level, marital status, regional mean household income, regional percentage of unemployment, and the difference of mean household income and percentage of unemployment, between parish and region.
